# Association of Domains of Sedentary Behavior with Cardiovascular Disease Events in The Multi-Ethnic Study of Atherosclerosis

**DOI:** 10.1101/2023.06.27.23291977

**Authors:** Nicole L. Mayo, Daniel A. Lopez, Robert C. Block, Hangchuan Shi, Alain G. Bertoni, Keith M. Diaz, Jingzhong Ding, Wendy S. Post, Yongmei Liu, Dongmei Li

**Affiliations:** Department of Public Health Sciences, Division of Epidemiology, University of Rochester Medical Center, Rochester, NY 14642; Department of Clinical & Translational Research, University of Rochester Medical Center, Rochester, NY 14642; Department of Epidemiology and Prevention, Wake Forest School of Medicine, Winston-Salem NC 27157; Department of Medicine, Division of Cardiology, Columbia University, New York, NY 10027; Department of Medicine, Wake Forest Baptist Medical Center, Winston-Salem, NC 27157; Department of Medicine, Division of Cardiology, Johns Hopkins University, School of Medicine, Baltimore, MD 21287; Department of Medicine, Division of Cardiology, Duke Molecular Physiology Institute, Duke University Medical Center, Durham, NC, 27701

**Keywords:** Mortality, Black, Chinese, Hispanic, Caucasian, Vascular

## Abstract

**Background:** Sedentary behavior is associated with an increased risk for adverse health outcomes, including cardiovascular disease (CVD), independent of physical activity status. Little is known about this relationship in an ethnically diverse population. The objective of our study is to assess the effects of leisure time and occupational sedentary behavior on multiple cardiovascular outcomes in a multi-ethnic cohort.

**Methods:** The Multi-Ethnic Study of Atherosclerosis (MESA) includes 2619 Caucasian, 1495 Hispanic, 1891 Black, and 804 Chinese-American adults aged 45-84 years and free of clinical CVD at enrollment, Sedentary behavior was self-reported at baseline. Participants were followed for an average of 13.6 years, and 14 types of cardiovascular outcomes were ascertained. Hazards of each cardiovascular outcome were modeled with adjustment for potential confounders, including physical activity.

**Results:** Every one hour per day increase in leisure time sedentary behavior predicts a 6% increase in the adjusted hazards for CVD death (*P* < 0.05). Every one hour increase in occupational sedentary time predicts a 21% and 20% decrease in the hazard for PVD and other revascularization, respectively (*P* < 0.05).

**Conclusions:** Leisure time sedentary behavior was associated with increased hazards for CVD death, but occupational sedentary time appears to be protective of peripheral vascular disease and other revascularization.

**Condensed Abstract:** Sedentary behavior has been consistently associated with an increased risk for adverse health outcomes, including cardiovascular disease (CVD), independent of physical activity status. The Multi-Ethnic Study of Atherosclerosis (MESA) consists of a racially and ethnically diverse cohort of adults age 45-84, free from CVD at baseline. Greater levels of leisure time sedentary behavior predicted increased hazards for PVD and CVD death after an average follow up of 13.6 years whereas occupational sedentary behaviors predicted reduced PVD. These results underscore the importance of reducing time spent sitting in addition to advocating for meeting physical activity targets across ethnicities.

## Introduction

Sedentary behavior is a distinct risk factor for a number of chronic conditions, and is potentially independent from moderate-vigorous intensity physical activity levels. A strong and consistent association has been observed between high levels of sedentary time and an increased risk for cardiovascular disease (CVD) mortality, all-cause mortality, incident type II diabetes, metabolic syndrome, depression, and some types of cancer (1-8). The National Health and Nutrition Examination Survey (NHANES) demonstrated that U.S. adults spend over half of their waking hours sedentary (9-11). Further, in 2018, adults 45 to 54 years old engaged in about 3.3 hours of leisure sedentary activities per day (12), which includes watching television, reading, relaxing, and using the computer for leisure.

Individuals who meet physical activity recommendations (13) can offset some of the deleterious effects of high sedentary time. Those who are inactive can improve their cardiometabolic profiles by replacing sedentary time with light or moderate activities, with greater benefits associated with replacing sedentary time with activities of higher intensities (14-16). Specifically, replacing 30 minutes of sedentary time per day with light activity can not only reduce mortality risk, but can also improve several cardiometabolic biomarkers. These effects are more pronounced if sedentary time is replaced with activities of greater intensities (16). However, in order to fully eliminate the increased mortality risk of eight or more hours of sedentary time per day, one must partake in at least 60-75 minutes of moderate activity each day, which may not be feasible for all (17). This highlights the importance of striving to reduce time spent sedentary, regardless of time spent active.

An increasing amount of evidence indicates that certain types of sedentary behavior may lead to greater CVD risk than others, which has led to growing conclusions that not all sedentary behavior is created equal (18, 19). Several studies have concluded that leisure time sedentary behavior, primarily television (TV) viewing, is more strongly associated with CVD risk factors (e.g., adiposity, C-reactive protein, fibrinogen, blood lipids) compared with non-leisure forms of sedentary behavior (e.g., occupational sedentary time (20-22). For example, among 7,660 middle-aged adults in the 1958 British Birth Cohort, higher levels of TV viewing, but not occupational sitting, had an adverse association with CVD biomarkers including C-reactive protein and fibrinogen (19). Another example is from a Danish population-based study in which 2,544 adults reported no significant associations between occupational sitting and cardiometabolic risk factors, but leisure time sitting was strongly associated with cardiometabolic risk factors including low-density lipoprotein cholesterol, cardiorespiratory fitness, and adiposity measures(23). Important gaps remain in the literature. First, many studies exploring sedentary behaviors and CVD have limited generalizability due to a lack of diverse representation. To our knowledge, the only study to date that examined sedentary behavior and CVD in an ethnically diverse population is the Multiethnic Cohort Study (MEC). The study sample had a racial/ethnic distribution of about 20-30% each for White, Japanese American, and Hispanic/Latino, followed by 15% for African-American and 7.5% for Native Hawaiian. The study observed an increase in all-cause and cardiovascular mortality for each additional unit increase in sedentary time (23). Second, existing studies documenting differential associations with CVD risk across domains of sedentary behavior have been largely cross-sectional in nature.

To help address these gaps, we examined the relationship between sedentary behavior and CVD outcomes in the Multi-Ethnic Study of Atherosclerosis (MESA). The objective of this study was to examine the relationship between leisure and non-leisure sedentary behavior and subsequent cardiovascular outcomes in this diverse cohort. Measures of sedentary behavior included a self-report of sedentary leisure-time, occupational sedentary behavior, and total sedentary time. Outcome measures included a longitudinal assessment of CVD events. We hypothesized that an increase in sedentary behavior was associated with an elevated risk of CVD, independent of physical activity. In addition, we hypothesized that the type of sedentary behavior (e.g., leisure vs occupational) was differentially associated with CVD outcomes.

## Methods

### Study Population

The present study utilizes data collected from MESA, which has been administered by the National Heart, Lung, and Blood Institute of the National Institutes of Health (24). Briefly, beginning in July 2000, 6814 men and women aged 45-84 years free from CVD were recruited from six locations in the United States (Baltimore City and Baltimore County, Maryland; Chicago, Illinois; Forsyth County, North Carolina; Los Angeles County, California; New York, New York; and St. Paul, Minnesota). Participants were recruited according to specific desired racial distributions of 38% non-Hispanic White, 28% African American, 23% Hispanic, and 11% Asian (predominantly Chinese). Following the first round of examinations from 2000-2002, five additional examinations have occurred (2002-2004, 2004-2005, 2005-2007, 2010-2011, 2016-2018) (24). In addition, yearly follow-up calls to participants collected information on several clinical outcomes including CVD, which were then verified from medical records by physicians. The current analysis utilized exposure and covariate data collected at the baseline examination and cardiovascular outcomes recorded through 2017.

### Assessment of Sedentary Behavior

Sedentary behaviors were assessed through an interviewer-administered MESA Typical Week Physical Activity Survey (TWPAS), which was designed to estimate habitual activities by asking participants about a typical week during the past month(25). The survey contains 28 question item categories including household chores, sports and dancing activities, care of others, transportation, walking pace, conditioning, lawn/yard/garden/farm, occupational activities, and leisure time activities. Participants were asked if they participated in certain activities during a typical week over the past month, then asked how much time per day (minutes) each activity was performed, and if the intensity was light, moderate, or vigorous. In order to assess leisure time sedentary activities, participants were asked: “In a typical week in the past month, did you sit or recline and watch TV?” and “In a typical week in the past month, did you read, knit, sew, visit, do nothing, or non-work recreational computer use?”. These items were recorded as single pursuits that were mutually exclusive (e.g., knitting while watching television was recorded as one category). After asking if participants worked to earn money, occupational sedentary time was assessed by the question: “At work, did you do light effort seated activities?”

### Assessment of Cardiovascular Outcomes

Cardiovascular events included myocardial infarction (MI), resuscitated cardiac arrest (RCA), angina pectoris (ANG), percutaneous transluminal coronary angioplasty (PTCA), coronary artery bypass graft (CABG), heart failure (HF), non-hemorrhagic stroke, transient ischemic attack (TIA), coronary heart disease (CHD), cardiovascular disease (CVD), peripheral vascular disease (PVD), coronary revascularization (REVC), other revascularization (any arterial revascularization of the aorta or arteries of the abdomen, pelvis, or legs), and cardiovascular disease death. Time to first event data were calculated as time from baseline to event. Each participant was contacted every 9-12 months to collect information on new CVD conditions, which were confirmed by a physician adjudication committee using death certificates and medical records (24).

### Assessment of covariates

Participants completed questionnaires to ascertain information regarding demographics, tobacco and alcohol use, medical conditions and access to medical care, family and reproductive history, and use of prescription medications. The covariates we considered for inclusion in our models are the demographic variables age, sex, race/ethnicity, education (less than high school, high school, some college, and college degree), and study site, as well as standard CVD risk factors such as smoking status (current, former/never), waist circumference, and Moderate to Vigorous Physical activity (MVPA) MET hours per day (derived from the TWPAS described above). MVPA MET hours per day is calculated as the sum of total moderate and vigorous MET minutes per week, converted into hours per day.

### Statistical Analysis

Total minutes of sedentary behavior per week was calculated by summing responses from three questions assessing time spent seated and watching TV, other seated leisure activities, and seated occupational tasks among those who work. This variable was modeled continuously as hours/day spent sedentary, and separately for hours/day spent in leisure-time and occupational sedentary behavior. The combined and domain specific variables were additionally divided into tertiles, with the first tertile representing those with the lowest amount of sedentary time and the third tertile representing those with the greatest amount of sedentary time.

Potential confounders identified from the literature included age, race/ethnicity, sex, study site, education, smoking status, waist circumference, and MVPA MET hours per week (26-28). Bivariate analyses assessed tertiles of sedentary time by each confounder, using Chi-squared statistics for binary and categorical variables, and general linear models for continuous variables. Similar methods were used to examine associations between these potential confounders and each cardiovascular outcome. We assessed and found no evidence of multicollinearity by examining the variance inflation factor (VIF) for all variables for each outcome (all VIF values < 5.0).

Separate Cox proportional hazard models were constructed to model baseline total sedentary hours/day, leisure time sedentary hours/day, and occupational sedentary hours/day with time to each (first) cardiovascular outcome. Model 1 was the crude (unadjusted) model. Model 2 adjusted for demographic variables and cardiovascular risk factors including age, race/ethnicity, sex, study site, education, smoking status, and waist circumference. Model 3 contains the variables in model two, with the addition of MVPA MET hours per day. Finally, each fully adjusted model was re-assessed using the sedentary tertile variables, with tertile one being the reference category.

Given that not all MESA participants have occupational sedentary hours to report (such as those that are unemployed or retired), we conducted sensitivity analyses that included only the working population for both leisure and occupational sedentary hours.

### Mediation analysis

Certain variables may be on the causal pathway between sedentary behavior and CVD. Therefore, we chose not to adjust for the variables: type II Diabetes, lipids (Total cholesterol, HDL-C, LDL-C, and triglycerides), and hypertension, and instead conducted mediation analyses using those variables as mediators. The mediation analyses were performed with the structural equation modeling (SEM) using Mplus version 8.5 (Muthen and Muthen, 2017). The model diagram was showed in Supplementary Table 4. Briefly, two regression models were fitted (i.e., mediator on sedentary time, and CVD outcome on mediator) on the basis of an original Cox proportional hazard model (i.e., CVD outcome on sedentary time). All the parameters were estimated by a maximum likelihood method and the confidence intervals were calculated using the Bootstrap method with the iteration number of 1,000.

## Results

We excluded 308 individuals who reported more than 24 hours of physical activity per day, leaving a sample of 6506. Participants were followed for an average of 13.6 years (SD=4.4 years). Mean age at baseline was 62.1 years (SD=10.2 years), and 52.9% of participants were female. 3047 (47%) of the population reported at baseline that they were currently employed. Participants spent on average 4.6 (SD=2.7) hours/day sedentary, with an average of 3.3 hours coming from leisure time and 2.4 hours coming from occupational sedentary behavior (for those employed). From 2000-2017 the number of adjudicated CVD events was 1044 CVD (all), 697 CHD (all), 425 REVC, 359 ANG, 385 HF, 364 CVD deaths, 336 MI, 304 PTCA, 246 Non-Hemorrhagic Stroke, 162 CABG, 122 PVD, 99 TIA, 84 other REVC, and 40 RCA.

**Table 1** displays population characteristics by tertile of overall sedentary time. With increasing tertile of sedentary hours per day, we observed increased waist circumference (T1: 96.7cm, T3: 99.8cm, p<0.0001) and decreased MVPA (T1: 14.5 hours/day, T3: 9.9 hours/day, p<0.0001).

**Table 1.** Baseline Characteristics of MESA Participants by Total Sedentary Behavior Tertiles.

|  | Total Sedentary Hours Per Day |  |  |  |
| --- | --- | --- | --- | --- |
|  | ≤3 h/d<br>(n=2209) | 3-5 h/d<br>(n=2145) | >5 h/d<br>(n=2152) | p value |
| Characteristic <sup>a</sup> |  |  |  |  |
| Age, years | 63.3±10.2 | 63.7±10.2 | 60.1±10.0 | <.0001 |
| Sex |  |  |  |  |
| Female | 1149(52.5) | 1159(54.1) | 1084(50.4) | 0.0547 |
| Male | 1040(47.5) | 984(45.9) | 1066(49.6) |  |
| Race/Ethnicity |  |  |  |  |
| White | 690(31.5) | 854(39.9) | 986(45.9) | <.0001 |
| Chinese American | 320(14.6) | 271(12.7) | 207(9.6) |  |
| Black, African American | 483(22.1) | 573(26.7) | 662(30.8) |  |
| Hispanic | 696(31.8) | 445(20.8) | 295(13.7) |  |
| Education |  |  |  |  |
| < High School | 634(29.0) | 350(16.4) | 208(9.7) | <.0001 |
| High School | 431(19.7) | 413(19.3) | 325(15.1) |  |
| Some College/Technical Certificate | 466(21.3) | 502(23.5) | 514(23.9) |  |
| College/Graduate Degree | 656(30) | 876(40.9) | 1101(51.3) |  |
| Smoking Status |  |  |  |  |
| Former/Never Smoker | 1952(89.2) | 1880(87.7) | 1818(84.6) | <.0001 |
| Current Smoker | 237(10.9) | 263(12.3) | 332(15.4) |  |
| Waist circumference (cm) | 96.7±13.6 | 97.6±14.1 | 99.8±15.2 | <.0001 |
| MVPA Met-hours per day | 14.5±13.1 | 12.2±10.4 | 9.9±8.5 | <.0001 |
| Study Site |  |  |  | <.0001 |
| Forsyth County, North Carolina | 272(12.4) | 361(16.9) | 384(17.9) |  |
| New York City, New York | 341(15.6) | 376(17.6) | 333(15.5) |  |
| Baltimore, Maryland | 280(12.8) | 334(15.6) | 362(16.8) |  |
| Twin Cities, Minnesota | 396(18.1) | 296(13.8) | 316(14.7) |  |
| Chicago, Illinois | 313(14.3) | 343(16.0) | 476(22.1) |  |
| Los Angeles, California | 587(26.8) | 433(20.2) | 279(13.0) |  |
| <sup>a</sup> Values are expressed as mean ± SD for continuous variables and N (%) for categorical variables<br>MVPA = Moderate to Vigorous Physical Activity |  |  |  |  |

When examined by hours per day (**Table 2**), for every one-hour increase in overall sedentary time, the hazards for CVD death increased by 3% (HR: 1.03, 95% CI: 0.99,1.08) after adjusting for demographics and CVD risk factors, including MVPA. This was predominately driven by leisure-time sedentary behavior, which was associated with a 6% (HR: 1.06, 95% CI: 1.00, 1.11) increased hazard for CVD death for every hour increase, where there was no significantly increased risk associated with occupational sedentary time. We also observed a reduced risk of PVD (HR: 0.79, 95% CI: 0.66,0.94), and other revascularization (HR: 0.80, 95% CI: 0.64,0.97) with increasing hours of occupational sedentary time in the fully adjusted models (**Supplemental Table 1**).

**Table 2.** Hazards of Cardiovascular Outcome by Sedentary Hours Per Day.

|  |  | Total Sedentary | Leisure Time | Occupational |
| --- | --- | --- | --- | --- |
| Cardiovascular Disease Death | Model 1 | 0.99(0.95,1.03) | 1.16(1.11,1.21) | 0.91(0.83,0.99) |
|  | Model 2 | 1.04(1.00,1.09) | 1.06(1.01,1.11) | 0.99(0.90,1.09) |
|  | Model 3 | 1.03(0.99,1.08) | 1.06(1.00,1.11) | 1.00(0.90,1.09) |
| Cardiovascular Disease (All) | Model 1 | 0.97(0.95,1.00) | 1.07(1.04,1.10) | 0.95(0.91,1.00) |
|  | Model 2 | 1.00(0.98,1.03) | 1.00(0.97,1.03) | 1.00(0.96,1.05) |
|  | Model 3 | 1.00(0.98,1.03) | 1.00(0.97,1.03) | 1.01(0.96,1.05) |
| Coronary Heart Disease (All) | Model 1 | 0.98(0.95,1.01) | 1.07(1.03,1.10) | 0.96(0.92,1.01) |
|  | Model 2 | 1.00(0.97,1.03) | 1.01(0.97,1.05) | 1.00(0.94,1.05) |
|  | Model 3 | 1.00(0.97,1.03) | 1.01(0.97,1.05) | 1.01(0.95,1.07) |
| Model 1 is the crude (unadjusted) model.<br>Model 2 is adjusted for age, sex, race, education, study site, smoking status, and waist circumference.<br>Model 3 is adjusted for variables in model 2 and Moderate to Vigorous Physical Activity (MVPA) |  |  |  |  |

**Table 3** displays the hazard ratios for tertiles of overall sedentary time and domains of sedentary time for our three primary outcomes. We did not observe significantly different hazards for any of our primary outcomes in our fully adjusted models for overall or domain specific sedentary time. One notable finding was that increasing tertile of occupational sedentary time was protective against PVD in the fully adjusted model (HR: 0.38, 95% CI: 0.15,0.91); **Supplemental Table 3**), similar to what we observed when modeled as hours per day. We explored interactions between sedentary time and age, race/ethnicity categories, and MVPA, and did not observe any significant interactions for adjusted models (p-interaction >0.10).

**Table 3.**
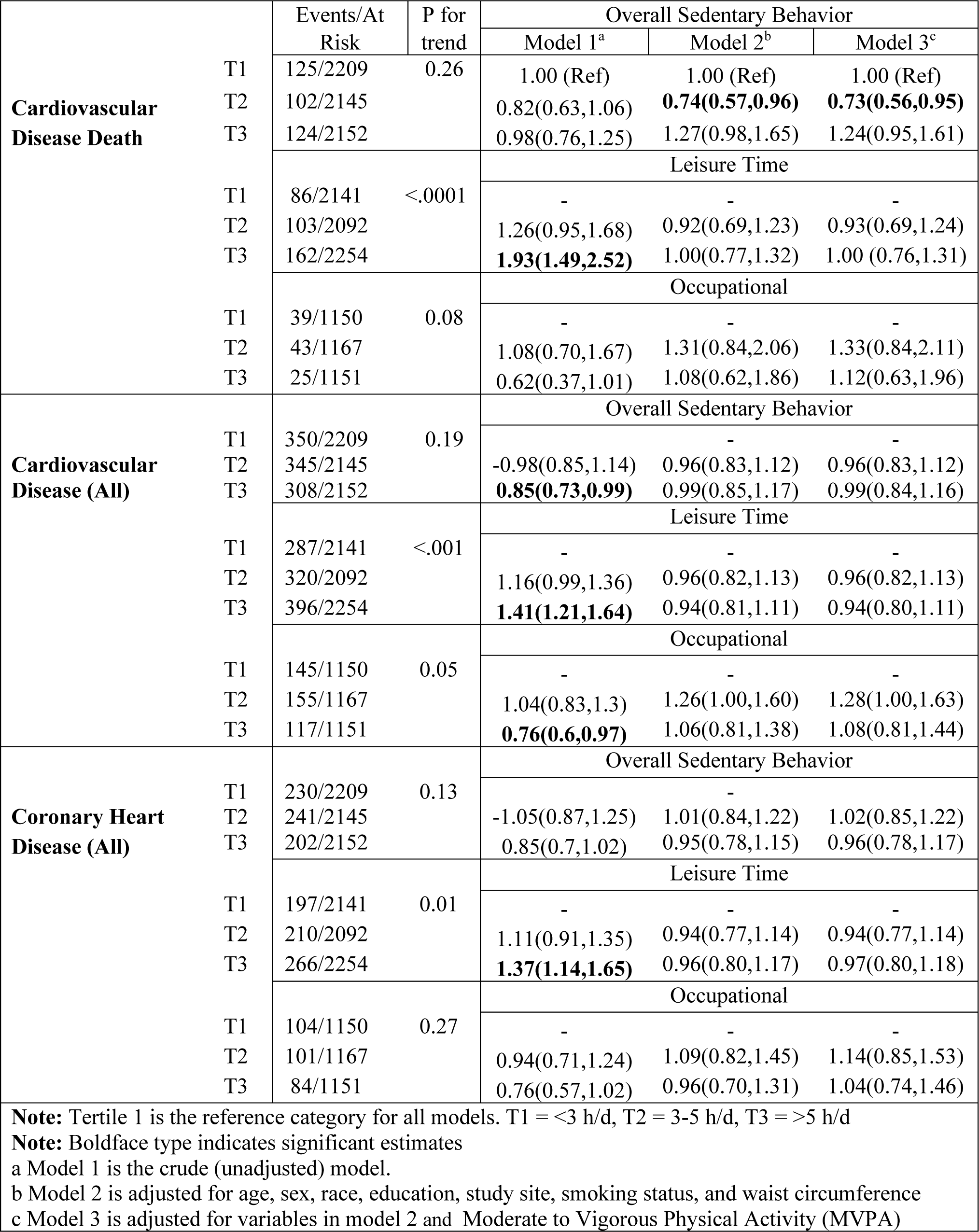
Hazards of Cardiovascular Outcome Type by Tertile of Sedentary Hours/day (HR (95% CI))

We conducted sensitivity analyses by restricting the sample to only those who were employed at baseline, in order to compare the results of the two different sedentary domains in the same population. We observed a similar protective effect of occupational sedentary time on PVD and other revascularization, but the results for CVD death were no longer significant.

Finally, the results of our mediation analyses (Supplementary Table 4) showed that hypertension, type II diabetes, and waist circumference all served as significant mediators in the association between CVD outcomes (i.e., CVD [all], CVD death, and CHD [all]) and the highest tertile of leisure-time sedentary behavior, while only hypertension and waist circumference served as mediators in the association between CVD outcomes and the middle tertile of leisure-time sedentary behavior. The positive dose-response phenomena were also observed for the mediation effects across the leisure-time sedentary behavior tertiles, indicating the growing contribution of hypertension, type II diabetes, and waist circumference on CVD outcomes when leisure sedentary time is increasing.

In contrast, compared with leisure sedentary time models, the mediation effects of all mediators were much lesser in occupational sedentary time models, and hypertension and type II diabetes did not serve as mediators in the association between CVD outcomes and the middle tertile of leisure-time sedentary behavior.

## Discussion

The most significant finding of our study was that the hazards for CVD event types varied depending on the domain of sedentary behavior examined. Increasing levels of leisure time sedentary behavior were associated with increased hazards for CVD death after an average follow-up of 13.6 years. Occupational sedentary time appears to be protective of vascular disease, including peripheral vascular disease and other vascularization. The four ethnicities in different geographic regions of the US, as well as the approximately 14 years longitudinal follow-up increases the causality of the study results. This study adds to the work done with the Multiethnic Cohort Study (23), by exploring associations in a more geographically diverse sample.

The findings that occupational sedentary time may be protective of certain cardiovascular event types has implications for sedentary behavior research. We observed associations between domain specific sedentary behavior and several cardiovascular event types that were not apparent when examining overall sedentary time. This is particularly important given that for certain cardiovascular events, leisure-time sedentary behavior was associated with increased hazards and occupational sedentary time was associated with decreased hazards. In addition, the mediation effects of hypertension, type II diabetes, and waist circumference were much smaller in the association of certain CVD outcome with occupational sedentary time, compared with the corresponding association with leisure sedentary time. The interpretation of these mediation analyses support that the effects of leisure-time sedentary time are less direct (mediated by hypertension, type II diabetes, and waist circumference) than they are for occupational sedentary time.

Additionally, the hazards of CVD death increased by 3% for every one hour increase in overall sedentary time, and 6% specifically for every hour increase in leisure time sedentary behavior, after adjusting for demographic variables and CVD risk factors, including MVPA. This is supported by a meta-analysis of 34 studies examining sedentary behavior (mostly through self-report) and CVD mortality, which observed 4% increased risk for each additional hour of daily sedentary time above six hours, and an 8% increased risk for each additional hour of daily TV time above 4 hours, after adjusting for physical activity (29). Additionally, Dunstan and colleagues observed an 18% increased hazard for CVD mortality for every one hour increase of TV time per day (30). Finally, a recent meta-analysis reported a 4% increased hazard for CVD mortality with each additional hour spent sitting per day and a 7% increased hazards for each additional hour of TV time (31). Our results now reveal this being present in a multi-ethnic cohort.

### Limitations

Even though there currently is not a “gold-standard” for measuring sedentary behaviors, it is still generally accepted that more objective methods such as accelerometers are less prone to measurement error compared to self-report (32). Sedentary time is systematically underreported (4, 33, 34), which could result in non-differential exposure misclassification and biased estimates towards the null. This could lead to the true effect of sedentary behavior on our outcomes being even greater than what we have observed. Occupational sedentary time was characterized as ‘light work while seated’, so it is possible that some of these seated occupational tasks may have required enough movement to raise energy expenditure levels into the category of light physical activity, which may explain the protective effects we observed. Unfortunately, specific occupation information was not available, so we were unable to examine differences by sedentary occupation. In addition, people with high social and economic statuses have more sedentary jobs than people with low social and economic statuses. Although we have controlled for education in our model, our study could miss some important confounding variables, such as the social determinants of health beyond education level, due to the lack of information in the data analyzed.

Further, some of the cardiovascular outcomes examined had relatively small sample sizes, which limited the power for identifying significant associations between sedentary behaviors and those outcomes. Finally, in our analyses we only assessed baseline sedentary behavior. As sedentary behavior has been shown to increase with age, further investigation is warranted for the potential time-varying effects of sedentary behavior on cardiovascular outcomes.

## Conclusions

In conclusion, greater levels of self-reported leisure time sedentary behavior are associated with increased hazards for CVD death, independent of MVPA. Occupational sedentary time appears to be protective against vascular disease. Care should be taken when examining overall sedentary time in relation to cardiovascular events due to the potential for mixing effects of different domains of sedentary behavior.

## Perspectives

Results of this study have implications for the 6 domains delineated by the Accreditation Council on Graduate Medical Education (ACGME) for medical providers. For example, a need exists for medical providers to carry-out recommendations for their patients of *diverse ethnic backgrounds* to reduce sedentary behavior. Given that reducing sedentary behavior lowers risk of CVD death, these recommendations are *appropriate and compassionate*, given that quality of life can also be enhanced. Our results add to current scientific evidence and *provider knowledge* which can guide them to be even more confident in their recommendations to their patients regarding the benefits of physical activity. These results can also support medical providers *improving their medical practice approaches* by assimilating the results into how their teams approach patients consistently regarding the importance of limiting sedentary behaviors. *Interpersonal and communication skills* used by providers to enhance patient vascular health can also be improved by considering these results. Finally, the results can enhance *systems-based practice* that medical providers work within.

Sedentary behavior has recently become a topic of considerable interest in The Physical Activity Guidelines for Americans (35). It addresses the risks of too much sitting for adults but does not prescribe a quantitative key guideline for sitting time or how to break-up sitting duration throughout the day. This is due to recent evidence demonstrating a complex relationship between the effects of sitting time and duration of moderate-to-vigorous physical activity on all-cause and cardiovascular disease mortality (36). With more moderate-to-vigorous physical activity, the risk of a given amount of sitting time is reduced. However, given the low amount of moderate-to-vigorous physical activity currently performed by most people in the United States, increasing physical activity and decreasing sitting are both likely to provide benefits. More research is warranted in order to help motivate and facilitate patients’ reduction of their sedentary time.

## Data Availability

The Multi-Ethnic Study of Atherosclerosis (MESA) is a study of the characteristics of subclinical cardiovascular disease (disease detected non-invasively before it has produced clinical signs and symptoms) and the risk factors that predict progression to clinically overt cardiovascular disease or progression of the subclinical disease. MESA researchers study a diverse, population-based sample of 6,814 asymptomatic men and women aged 45-84. Approximately 38 percent of the recruited participants are white, 28 percent African-American, 22 percent Hispanic, and 12 percent Asian, predominantly of Chinese descent. MESA Coordinating Center distributes only deidentified datasets (with HIPAA defined identifiers removed) to Recipient Investigators with approved manuscript proposals in compliance with MESA and NHLBI/NIH data privacy and sharing standard practices and policy.

## Acknowledgments

None

## Abbreviations

ANG: Angina pectoris
CABG: Coronary artery bypass graft
CHD: Coronary heart disease
HF: Heart failure
CVD: Cardiovascular disease
MI: Myocardial infarction
PTCA: Percutaneous transluminal coronary angioplasty
PVD: Peripheral vascular disease
RCA: Resuscitated cardiac arrest
TIA: Transient ischemic attack
MVPA: Moderate to Vigorous Physical Activity

---

**Supplemental Table 1.** Hazards of Cardiovascular Outcome by Sedentary Hours Per Day.

|  |  |  |  |  |
| --- | --- | --- | --- | --- |
|  |  | Total Sedentary | Leisure Time | Occupational |
| Myocardial Infarction | Model 1 | 0.97(0.93,1.01) | 1.05(1.00,1.11) | 0.94(0.88,1.02) |
|  | Model 2 | 0.99(0.95,1.04) | 1.01(0.95,1.06) | 0.99(0.91,1.06) |
|  | Model 3 | 1.00(0.95,1.04) | 1.01(0.95,1.06) | 1.01(0.92,1.09) |
| Heart Failure | Model 1 | 1.00(0.96,1.04) | 1.13(1.08,1.18) | 0.96(0.89,1.03) |
|  | Model 2 | 1.03(0.98,1.07) | 1.03(0.98,1.08) | 1.02(0.94,1.10) |
|  | Model 3 | 1.03(0.98,1.07) | 1.03(0.98,1.08) | 1.02(0.94,1.11) |
| Resuscitated Cardiac Arrest | Model 1 | 1.02(0.91,1.14) | 1.07(0.92,1.22) | 1.07(0.88,1.28) |
|  | Model 2 | 1.01(0.89,1.14) | 0.98(0.83,1.13) | 1.08(0.88,1.31) |
|  | Model 3 | 1.00(0.88,1.12) | 0.97(0.83,1.12) | 1.04(0.83,1.29) |
| Angina | Model 1 | 0.98(0.94,1.02) | 1.04(0.99,1.09) | 0.97(0.90,1.03) |
|  | Model 2 | 0.99(0.95,1.03) | 0.99(0.94,1.05) | 1.00(0.93,1.07) |
|  | Model 3 | 1.00(0.96,1.04) | 1.00(0.94,1.05) | 1.03(0.95,1.10) |
| PTCA | Model 1 | 0.98(0.94,1.02) | 0.98(0.92,1.03) | 1.00(0.93,1.06) |
|  | Model 2 | 0.97(0.93,1.02) | 0.95(0.89,1.01) | 0.99(0.92,1.06) |
|  | Model 3 | 0.97(0.93,1.02) | 0.95(0.90,1.01) | 1.01(0.93,1.09) |
| Coronary Bypass Graft | Model 1 | 0.97(0.91,1.03) | 1.03(0.95,1.10) | 0.96(0.87,1.06) |
|  | Model 2 | 0.96(0.90,1.02) | 0.99(0.91,1.07) | 0.98(0.88,1.09) |
|  | Model 3 | 0.97(0.90,1.03) | 0.99(0.91,1.07) | 1.00(0.89,1.12) |
| Peripheral Vascular Disease | Model 1 | 0.98(0.92,1.05) | 1.15(1.07,1.24) | **0.78(0.66,0.91)** |
|  | Model 2 | 0.99(0.92,1.06) | 1.07(0.98,1.16) | **0.80(0.66,0.93)** |
|  | Model 3 | 0.99(0.92,1.07) | 1.07(0.98,1.16) | **0.79(0.66,0.94)** |
| Stroke | Model 1 | 0.96(0.92,1.01) | 1.07(1.01,1.13) | 0.92(0.84,1.01) |
|  | Model 2 | 1.00(0.95,1.06) | 1.00(0.93,1.06) | 0.99(0.89,1.08) |
|  | Model 3 | 1.00(0.94,1.05) | 0.99(0.93,1.06) | 0.97(0.87,1.07) |
| Transient Ischemic Attack | Model 1 | 0.98(0.91,1.06) | 1.09(1.00,1.19) | 0.88(0.76,1.01) |
|  | Model 2 | 1.01(0.93,1.09) | 1.02(0.93,1.13) | 0.96(0.82,1.10) |
|  | Model 3 | 1.02(0.94,1.10) | 1.03(0.93,1.13) | 1.00(0.85,1.16) |
| Coronary Revascularization | Model 1 | 0.97(0.94,1.01) | 0.99(0.94,1.04) | 0.99(0.93,1.05) |
|  | Model 2 | 0.97(0.93,1.00) | 0.96(0.91,1.01) | 1.00(0.93,1.06) |
|  | Model 3 | 0.97(0.93,1.01) | 0.96(0.91,1.01) | 1.01(0.94,1.07) |
| Other Revascularization | Model 1 | 0.92(0.84,1.00) | 1.06(0.96,1.17) | **0.73(0.60,0.87)** |
|  | Model 2 | 0.92(0.84,1.01) | 0.98(0.88,1.09) | **0.77(0.63,0.93)** |
|  | Model 3 | 0.93(0.85,1.03) | 0.99(0.89,1.10) | **0.80(0.64,0.97)** |
| **Note:** Boldface type indicates significant estimates<br>Model 1 is the crude (unadjusted) model.<br>Model 2 is adjusted for age, sex, race, education, study site, smoking status, and waist circumference.<br>Model 3 is adjusted for variables in model 2 and Moderate to Vigorous Physical Activity (MVPA) |  |  |  |  |

**Supplemental Table 2.** Hazards of Cardiovascular Outcome Type by Tertile of Total Sedentary Hours.

|  |  |  |  |  |  |  |
| --- | --- | --- | --- | --- | --- | --- |
|  |  | Events/At Risk | P for trend | Model 1<sup>a</sup><br>HR (95% CI) | Model 2<sup>b</sup><br>HR (95% CI) | Model 3<sup>c</sup><br>HR (95% CI) |
| Myocardial Infarction | T1 | 114/2209 | 0.09 | 1.00 (Ref) | 1.00 (Ref) | 1.00 (Ref) |
|  | T2 | 121/2145 |  | 1.06(0.82,1.37) | 1.08(0.83,1.40) | 1.09(0.84,1.41) |
|  | T3 | 91/2152 |  | 0.77(0.58,1.02) | 0.92(0.69,1.22) | 0.93(0.69,1.24) |
| Heart Failure | T1 | 121/2209 | 0.83 | - | - | - |
|  | T2 | 125/2145 |  | 1.03(0.81,1.33) | 0.96(0.74,1.23) | 0.96(0.74,1.24) |
|  | T3 | 128/2152 |  | 1.03(0.80,1.32) | 1.16(0.89,1.51) | 1.16(0.89,1.51) |
| Resuscitated Cardiac Arrest | T1 | 13/2209 | 0.84* | - | - | - |
|  | T2 | 12/2145 |  | 0.92(0.42,2.03) | 0.81(0.36,1.80) | 0.79(0.35,1.75) |
|  | T3 | 15/2152 |  | 1.13(0.53,2.40) | 1.04(0.48,2.27) | 0.94(0.43,2.10) |
| Angina | T1 | 112/2209 | 0.09 | - | - | - |
|  | T2 | 131/2145 |  | 1.18(0.92,1.52) | 1.16(0.90,1.50) | 0.94(0.43,2.10) |
|  | T3 | 100/2152 |  | 0.88(0.67,1.15) | 0.92(0.70,1.23) | 1.18(0.91,1.53) |
| PTCA | T1 | 101/2209 | 0.51 | - | - | - |
|  | T2 | 104/2145 |  | 1.03(0.78,1.35) | 1.01(0.76,1.33) | 1.01(0.77,1.34) |
|  | T3 | 89/2152 |  | 0.86(0.64,1.14) | 0.81(0.60,1.09) | 0.82(0.60,1.12) |
| Coronary Bypass Graft | T1 | 54/2209 | 0.76 | - | - | - |
|  | T2 | 55/2145 |  | 1.02(0.70,1.49) | 0.96(0.66,1.40) | 0.98(0.67,1.43) |
|  | T3 | 48/2152 |  | 0.87(0.59,1.29) | 0.84(0.56,1.26) | 0.88(0.58,1.33) |
| Peripheral Vascular Disease | T1 | 39/2209 | 0.95 | - | - | - |
|  | T2 | 37/2145 |  | 0.95(0.60,1.49) | 0.88(0.56,1.39) | 0.89(0.56,1.41) |
|  | T3 | 40/2152 |  | 1.01(0.65,1.57) | 1.04(0.65,1.65) | 1.05(0.66,1.68) |
| Stroke | T1 | 76/2209 | 0.06 | - | - | - |
|  | T2 | 92/2145 |  | 1.20(0.89,1.63) | 1.19(0.87,1.62) | 1.18(0.86,1.60) |
|  | T3 | 64/2152 |  | 0.82(0.58,1.14) | 1.03(0.72,1.45) | 1.00(0.70,1.41) |
| Transient Ischemic Attack | T1 | 34/2209 | 0.74 | - | - | - |
|  | T2 | 33/2145 |  | 0.97(0.60,1.57) | 0.94(0.58,1.53) | 0.96(0.59,1.56) |
|  | T3 | 28/2152 |  | 0.81(0.49,1.33) | 0.92(0.54,1.54) | 0.96(0.56,1.62) |
| Coronary Revascularization | T1 | 145/2209 | 0.34 | - | - | - |
|  | T2 | 143/2145 |  | 0.99(0.78,1.25) | 0.95(0.75,1.20) | 0.96(0.76,1.21) |
|  | T3 | 123/2152 |  | 0.83(0.65,1.05) | 0.79(0.61,1.02) | 0.81(0.62,1.04) |
| Other Revascularization | T1 | 37/2209 | 0.09 | - | - | - |
|  | T2 | 23/2145 |  | 0.62(0.36,1.03) | **0.59(0.34,0.99)** | 0.61(0.35,1.02) |
|  | T3 | 22/2152 |  | **0.58(0.34,0.98)** | 0.60(0.34,1.03) | 0.64(0.36,1.12) |
| **Note:** Tertile 1 is the reference category for all models. |  |  |  |  |  |  |
| **Note:** Boldface type indicates significant estimates |  |  |  |  |  |  |
| a Model 1 is the crude (unadjusted) model. |  |  |  |  |  |  |
| b Model 2 is adjusted for age, sex, race, education, study site, smoking status, and waist circumference |  |  |  |  |  |  |
| c Model 3 is adjusted for variables in model 2 and Moderate to Vigorous Physical Activity (MVPA) |  |  |  |  |  |  |

**Supplemental Table 3.**
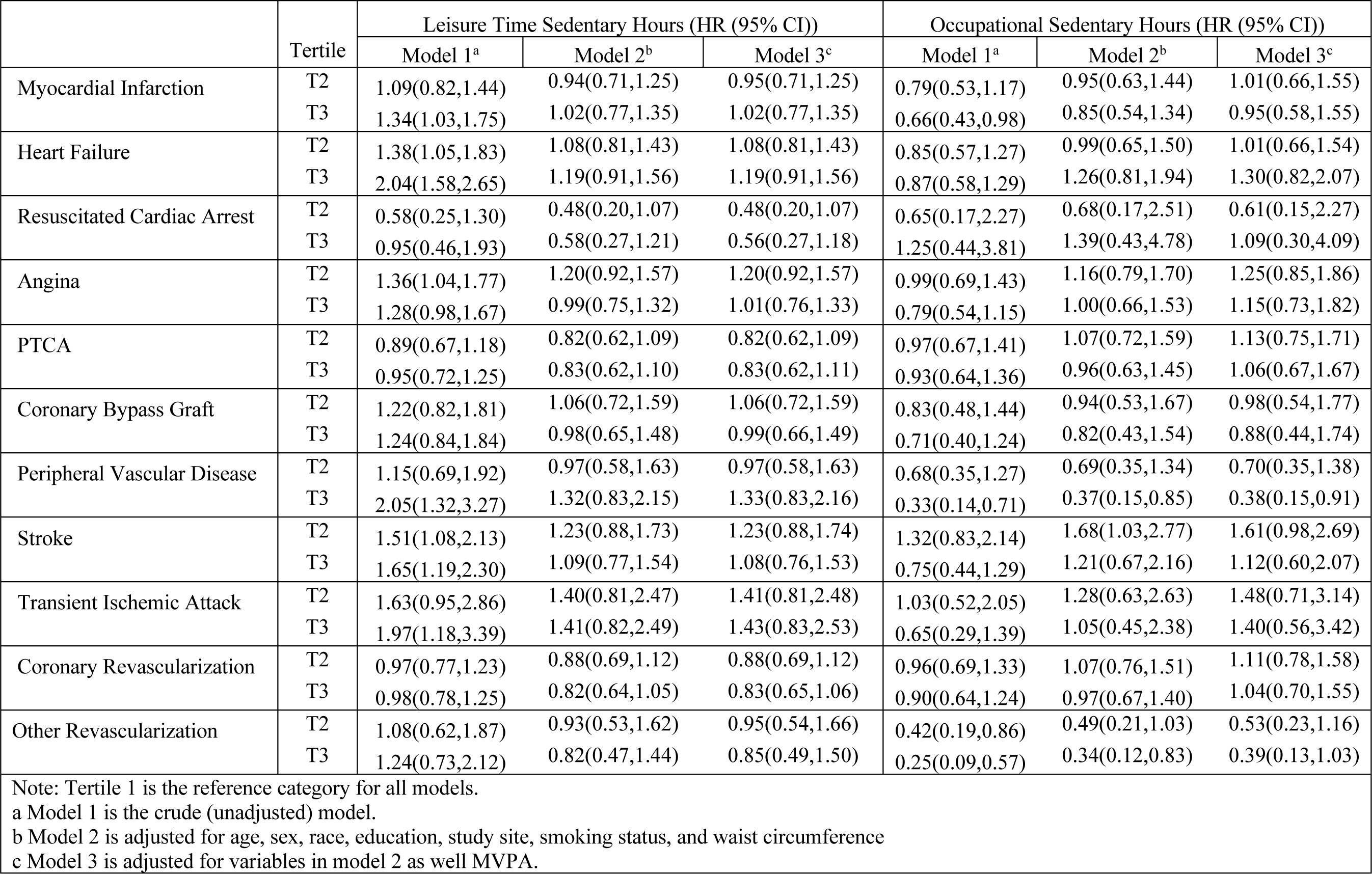
Hazards of Cardiovascular Outcome Type by Tertile of Leisure and Occupational Sedentary Hours.

|  | Tertile | Leisure Time Sedentary Hours (HR (95% CI)) |  |  | Occupational Sedentary Hours (HR (95% CI)) |  |  |
| --- | --- | --- | --- | --- | --- | --- | --- |
|  |  | Model 1<sup>a</sup> | Model 2<sup>b</sup> | Model 3<sup>c</sup> | Model 1<sup>a</sup> | Model 2<sup>b</sup> | Model 3<sup>c</sup> |
| Myocardial Infarction | T2 | 1.09(0.82,1.44) | 0.94(0.71,1.25) | 0.95(0.71,1.25) | 0.79(0.53,1.17) | 0.95(0.63,1.44) | 1.01(0.66,1.55) |
|  | T3 | 1.34(1.03,1.75) | 1.02(0.77,1.35) | 1.02(0.77,1.35) | 0.66(0.43,0.98) | 0.85(0.54,1.34) | 0.95(0.58,1.55) |
| Heart Failure | T2 | 1.38(1.05,1.83) | 1.08(0.81,1.43) | 1.08(0.81,1.43) | 0.85(0.57,1.27) | 0.99(0.65,1.50) | 1.01(0.66,1.54) |
|  | T3 | 2.04(1.58,2.65) | 1.19(0.91,1.56) | 1.19(0.91,1.56) | 0.87(0.58,1.29) | 1.26(0.81,1.94) | 1.30(0.82,2.07) |
| Resuscitated Cardiac Arrest | T2 | 0.58(0.25,1.30) | 0.48(0.20,1.07) | 0.48(0.20,1.07) | 0.65(0.17,2.27) | 0.68(0.17,2.51) | 0.61(0.15,2.27) |
|  | T3 | 0.95(0.46,1.93) | 0.58(0.27,1.21) | 0.56(0.27,1.18) | 1.25(0.44,3.81) | 1.39(0.43,4.78) | 1.09(0.30,4.09) |
| Angina | T2 | 1.36(1.04,1.77) | 1.20(0.92,1.57) | 1.20(0.92,1.57) | 0.99(0.69,1.43) | 1.16(0.79,1.70) | 1.25(0.85,1.86) |
|  | T3 | 1.28(0.98,1.67) | 0.99(0.75,1.32) | 1.01(0.76,1.33) | 0.79(0.54,1.15) | 1.00(0.66,1.53) | 1.15(0.73,1.82) |
| PTCA | T2 | 0.89(0.67,1.18) | 0.82(0.62,1.09) | 0.82(0.62,1.09) | 0.97(0.67,1.41) | 1.07(0.72,1.59) | 1.13(0.75,1.71) |
|  | T3 | 0.95(0.72,1.25) | 0.83(0.62,1.10) | 0.83(0.62,1.11) | 0.93(0.64,1.36) | 0.96(0.63,1.45) | 1.06(0.67,1.67) |
| Coronary Bypass Graft | T2 | 1.22(0.82,1.81) | 1.06(0.72,1.59) | 1.06(0.72,1.59) | 0.83(0.48,1.44) | 0.94(0.53,1.67) | 0.98(0.54,1.77) |
|  | T3 | 1.24(0.84,1.84) | 0.98(0.65,1.48) | 0.99(0.66,1.49) | 0.71(0.40,1.24) | 0.82(0.43,1.54) | 0.88(0.44,1.74) |
| Peripheral Vascular Disease | T2 | 1.15(0.69,1.92) | 0.97(0.58,1.63) | 0.97(0.58,1.63) | 0.68(0.35,1.27) | 0.69(0.35,1.34) | 0.70(0.35,1.38) |
|  | T3 | 2.05(1.32,3.27) | 1.32(0.83,2.15) | 1.33(0.83,2.16) | 0.33(0.14,0.71) | 0.37(0.15,0.85) | 0.38(0.15,0.91) |
| Stroke | T2 | 1.51(1.08,2.13) | 1.23(0.88,1.73) | 1.23(0.88,1.74) | 1.32(0.83,2.14) | 1.68(1.03,2.77) | 1.61(0.98,2.69) |
|  | T3 | 1.65(1.19,2.30) | 1.09(0.77,1.54) | 1.08(0.76,1.53) | 0.75(0.44,1.29) | 1.21(0.67,2.16) | 1.12(0.60,2.07) |
| Transient Ischemic Attack | T2 | 1.63(0.95,2.86) | 1.40(0.81,2.47) | 1.41(0.81,2.48) | 1.03(0.52,2.05) | 1.28(0.63,2.63) | 1.48(0.71,3.14) |
|  | T3 | 1.97(1.18,3.39) | 1.41(0.82,2.49) | 1.43(0.83,2.53) | 0.65(0.29,1.39) | 1.05(0.45,2.38) | 1.40(0.56,3.42) |
| Coronary Revascularization | T2 | 0.97(0.77,1.23) | 0.88(0.69,1.12) | 0.88(0.69,1.12) | 0.96(0.69,1.33) | 1.07(0.76,1.51) | 1.11(0.78,1.58) |
|  | T3 | 0.98(0.78,1.25) | 0.82(0.64,1.05) | 0.83(0.65,1.06) | 0.90(0.64,1.24) | 0.97(0.67,1.40) | 1.04(0.70,1.55) |
| Other Revascularization | T2 | 1.08(0.62,1.87) | 0.93(0.53,1.62) | 0.95(0.54,1.66) | 0.42(0.19,0.86) | 0.49(0.21,1.03) | 0.53(0.23,1.16) |
|  | T3 | 1.24(0.73,2.12) | 0.82(0.47,1.44) | 0.85(0.49,1.50) | 0.25(0.09,0.57) | 0.34(0.12,0.83) | 0.39(0.13,1.03) |

Note: Tertile 1 is the reference category for all models.
a Model 1 is the crude (unadjusted) model.
b Model 2 is adjusted for age, sex, race, education, study site, smoking status, and waist circumference
c Model 3 is adjusted for variables in model 2 as well MVPA.

**Supplemental Table 4.**
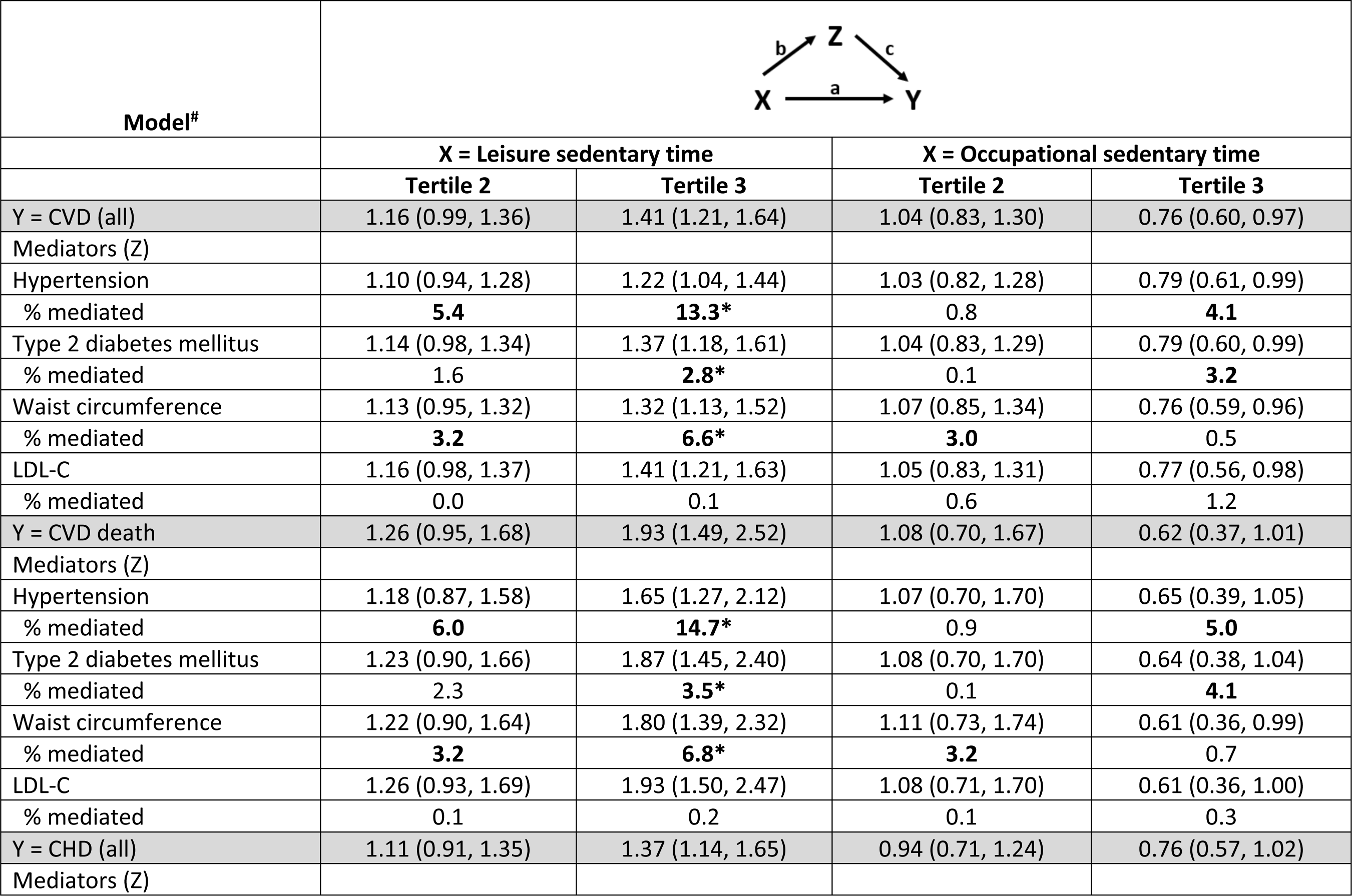
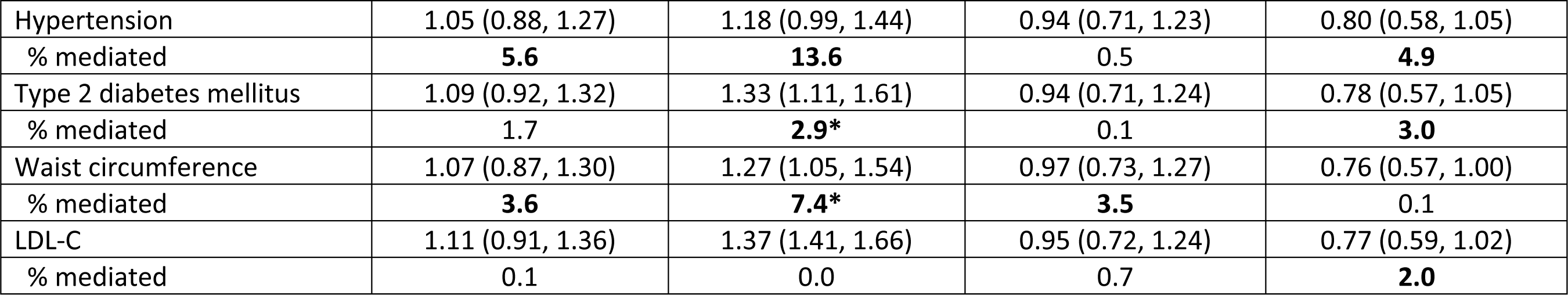
**Mediation analyses using hypertension, type 2 diabetes mellitus, waist circumference, and LDL as mediators.**

**Supplemental Table 5.**
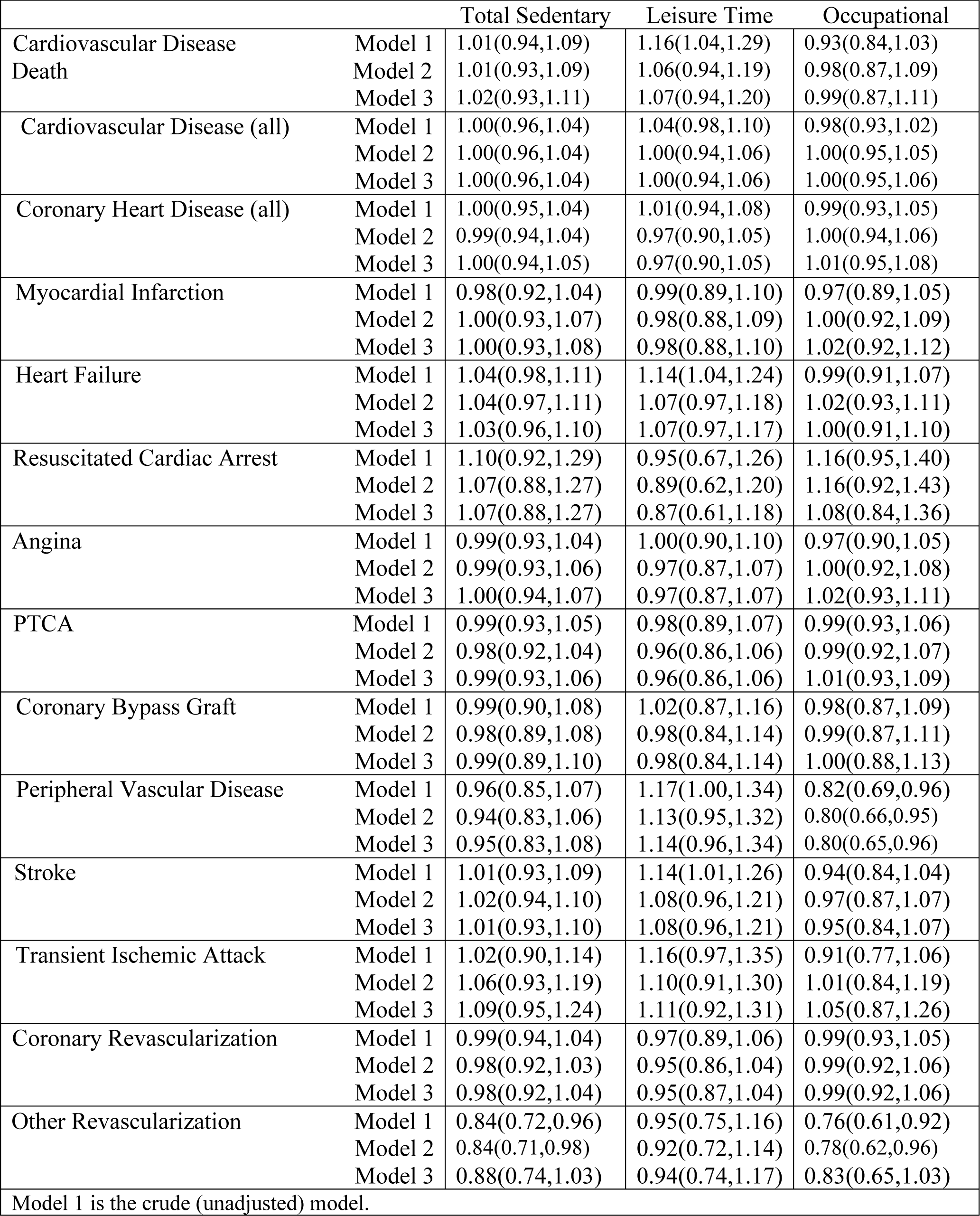
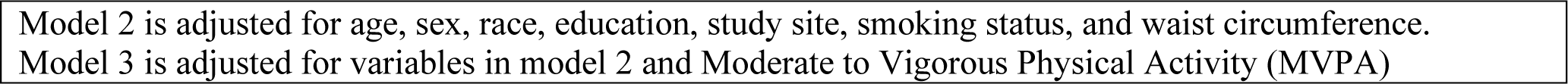
Hazards of Cardiovascular Outcome by Sedentary Hours Per Day Among those Employed at baseline (N=3047)

|  |  |  |  |  |
| --- | --- | --- | --- | --- |
|  |  | Total Sedentary | Leisure Time | Occupational |
| Cardiovascular Disease Death | Model 1 | 1.01(0.94,1.09) | 1.16(1.04,1.29) | 0.93(0.84,1.03) |
|  | Model 2 | 1.01(0.93,1.09) | 1.06(0.94,1.19) | 0.98(0.87,1.09) |
|  | Model 3 | 1.02(0.93,1.11) | 1.07(0.94,1.20) | 0.99(0.87,1.11) |
| Cardiovascular Disease (all) | Model 1 | 1.00(0.96,1.04) | 1.04(0.98,1.10) | 0.98(0.93,1.02) |
|  | Model 2 | 1.00(0.96,1.04) | 1.00(0.94,1.06) | 1.00(0.95,1.05) |
|  | Model 3 | 1.00(0.96,1.04) | 1.00(0.94,1.06) | 1.00(0.95,1.06) |
| Coronary Heart Disease (all) | Model 1 | 1.00(0.95,1.04) | 1.01(0.94,1.08) | 0.99(0.93,1.05) |
|  | Model 2 | 0.99(0.94,1.04) | 0.97(0.90,1.05) | 1.00(0.94,1.06) |
|  | Model 3 | 1.00(0.94,1.05) | 0.97(0.90,1.05) | 1.01(0.95,1.08) |
| Myocardial Infarction | Model 1 | 0.98(0.92,1.04) | 0.99(0.89,1.10) | 0.97(0.89,1.05) |
|  | Model 2 | 1.00(0.93,1.07) | 0.98(0.88,1.09) | 1.00(0.92,1.09) |
|  | Model 3 | 1.00(0.93,1.08) | 0.98(0.88,1.10) | 1.02(0.92,1.12) |
| Heart Failure | Model 1 | 1.04(0.98,1.11) | 1.14(1.04,1.24) | 0.99(0.91,1.07) |
|  | Model 2 | 1.04(0.97,1.11) | 1.07(0.97,1.18) | 1.02(0.93,1.11) |
|  | Model 3 | 1.03(0.96,1.10) | 1.07(0.97,1.17) | 1.00(0.91,1.10) |
| Resuscitated Cardiac Arrest | Model 1 | 1.10(0.92,1.29) | 0.95(0.67,1.26) | 1.16(0.95,1.40) |
|  | Model 2 | 1.07(0.88,1.27) | 0.89(0.62,1.20) | 1.16(0.92,1.43) |
|  | Model 3 | 1.07(0.88,1.27) | 0.87(0.61,1.18) | 1.08(0.84,1.36) |
| Angina | Model 1 | 0.99(0.93,1.04) | 1.00(0.90,1.10) | 0.97(0.90,1.05) |
|  | Model 2 | 0.99(0.93,1.06) | 0.97(0.87,1.07) | 1.00(0.92,1.08) |
|  | Model 3 | 1.00(0.94,1.07) | 0.97(0.87,1.07) | 1.02(0.93,1.11) |
| PTCA | Model 1 | 0.99(0.93,1.05) | 0.98(0.89,1.07) | 0.99(0.93,1.06) |
|  | Model 2 | 0.98(0.92,1.04) | 0.96(0.86,1.06) | 0.99(0.92,1.07) |
|  | Model 3 | 0.99(0.93,1.06) | 0.96(0.86,1.06) | 1.01(0.93,1.09) |
| Coronary Bypass Graft | Model 1 | 0.99(0.90,1.08) | 1.02(0.87,1.16) | 0.98(0.87,1.09) |
|  | Model 2 | 0.98(0.89,1.08) | 0.98(0.84,1.14) | 0.99(0.87,1.11) |
|  | Model 3 | 0.99(0.89,1.10) | 0.98(0.84,1.14) | 1.00(0.88,1.13) |
| Peripheral Vascular Disease | Model 1 | 0.96(0.85,1.07) | 1.17(1.00,1.34) | 0.82(0.69,0.96) |
|  | Model 2 | 0.94(0.83,1.06) | 1.13(0.95,1.32) | 0.80(0.66,0.95) |
|  | Model 3 | 0.95(0.83,1.08) | 1.14(0.96,1.34) | 0.80(0.65,0.96) |
| Stroke | Model 1 | 1.01(0.93,1.09) | 1.14(1.01,1.26) | 0.94(0.84,1.04) |
|  | Model 2 | 1.02(0.94,1.10) | 1.08(0.96,1.21) | 0.97(0.87,1.07) |
|  | Model 3 | 1.01(0.93,1.10) | 1.08(0.96,1.21) | 0.95(0.84,1.07) |
| Transient Ischemic Attack | Model 1 | 1.02(0.90,1.14) | 1.16(0.97,1.35) | 0.91(0.77,1.06) |
|  | Model 2 | 1.06(0.93,1.19) | 1.10(0.91,1.30) | 1.01(0.84,1.19) |
|  | Model 3 | 1.09(0.95,1.24) | 1.11(0.92,1.31) | 1.05(0.87,1.26) |
| Coronary Revascularization | Model 1 | 0.99(0.94,1.04) | 0.97(0.89,1.06) | 0.99(0.93,1.05) |
|  | Model 2 | 0.98(0.92,1.03) | 0.95(0.86,1.04) | 0.99(0.92,1.06) |
|  | Model 3 | 0.98(0.92,1.04) | 0.95(0.87,1.04) | 0.99(0.92,1.06) |
| Other Revascularization | Model 1 | 0.84(0.72,0.96) | 0.95(0.75,1.16) | 0.76(0.61,0.92) |
|  | Model 2 | 0.84(0.71,0.98) | 0.92(0.72,1.14) | 0.78(0.62,0.96) |
|  | Model 3 | 0.88(0.74,1.03) | 0.94(0.74,1.17) | 0.83(0.65,1.03) |
| Model 1 is the crude (unadjusted) model. |  |  |  |  |

Model 2 is adjusted for age, sex, race, education, study site, smoking status, and waist circumference. Model 3 is adjusted for variables in model 2 and Moderate to Vigorous Physical Activity (MVPA)

